# Age-associated B-cells are expanded in early arthritis linked to atherosclerosis and immune circuits - a potential role as a biomarker for risk stratification

**DOI:** 10.1101/2025.01.14.25320531

**Authors:** Daniel Miranda-Prieto, Mercedes Alperi-López, Ángel I. Pérez-Álvarez, Sara Alonso-Castro, Ana Suárez, Javier Rodríguez-Carrio

**Author notes:** **Corresponding authors:**Javier Rodríguez-Carrio, MSc, PhD Area of Immunology, Department of Functional Biology, Faculty of Medicine, University of Oviedo Campus El Cristo C/ Julián Clavería s/n, L-19 33006 – Oviedo Spain Prof. Ana Suárez, PhD Area of Immunology, Department of Functional Biology, Faculty of Medicine, University of Oviedo Campus El Cristo C/ Julián Clavería s/n, L-19 33006 – Oviedo Spain.

## Abstract

**Objective:** immune dysregulation may play a role in cardiovascular (CV) risk excess in rheumatoid arthritis (RA). However, exact mediators are unknown. Age-associated B cells (ABCs) have emerged as multi-faceted pro-inflammatory mediators, also in the atherosclerosis microenvironment, but their role in autoimmunity is ill-defined. Our aim was to evaluate ABCs levels in the earliest stages of inflammatory arthritis and their potential role as biomarkers of atherosclerosis.

**Methods:** ABCs were quantified by flow cytometry in 58 early RA patients, 11 individuals with clinical-suspect arthralgia (CSA) and 33 healthy controls (HC). Atherosclerosis occurrence was measured by Doppler-ultrasound. Cytokines were measured by multiplex immunoassays. Cardiometabolic-related proteins were evaluated using high-throughput targeted proteomics.

**Results:** Circulating ABCs were increased in RA patients compared with HC within the CD19+ and PBMCs pools (p=0.013 and p<0.001, respectively). Numerically higher ABCs levels were found in CSA. ABCs frequency was unrelated to disease features and traditional CV risk factors but negatively associated with good therapeutic outcomes upon csDMARD at 6 and 12 months. ABCs frequency was positively correlated with proinflammatory cytokines (IFNg, TNF, IL-6 and IL-21) and proteomic signatures related to B- and T-cell responses as well as cellular pathways linked to atherosclerosis.

ABCs were independently associated with atherosclerosis occurrence and extent in RA patients. Furthermore, adding ABCs levels significantly improved risk stratification over conventional instruments.

**Conclusions:** ABCs expansion is an early event along arthritis course, linked to therapeutic outcomes, inflammatory milieu and atherosclerosis burden. ABCs may be a missing link between humoral responses and atherosclerosis in autoimmunity.

## INTRODUCTION

Cardiovascular disease (CVD) burden is increased in rheumatoid arthritis (RA), although risk excess cannot be solely attributed to traditional CV risk factors [1]. Non-traditional CV risk factors such as chronic inflammation or autoimmune phenomena, are thought to play a role. A number of auto-antibodies have been described in this setting [2], althought the characterization of humoral responses beyond antibody production has received little attention. Importantly, the role of B-cells in RA is more complex than initially conceived, since targeted therapies have been demonstrated to abrogate B-cell responses to different degrees, and changes in autoantibody levels did not parallel those of clinical improvement [3,4]. These findings point to other roles of B-cells and highlight the need of a deeper study of the B-cell compartment.

Age-associated B-cells (ABCs) are a rare subset of B-cells [5,6]. Human ABCs exhibit a memory-like B-cell phenotype, and they are characterized by the expression of T-bet, CD11c, CD11b and lack of CD21 [7]. ABCs may be related to the atypical, double-negative 2 (DN2) B-cells, but evidence suggests they form an independent subset [7]. ABCs differentiate from naïve B-cells via a combination of B-cell receptor (BCR) ligation, engagement of toll-like receptors 7 or 9 (TLR-7/9) and cytokine signals (mostly IFN-γ and IL-21), either by cognate T-cells or bystander production [8,9]. ABCs may differentiate into plasma cells and produce cytokines and antibodies. Furthermore, ABCs can present antigens to T-cells due to elevated levels of MHC-II and CD11c, costimulatory signals, cytoskeletal rearrangement and higher expression of vesicular transport genes [5]. ABCs appear to show a unique functional program among other B-cell subsets, characterized by high expression of cytokines and their receptors, chemokines and adhesion molecules, as well as antigen presentation machinery and costimulatory molecules [5,9]. Collectively, these activities may lead to higher T-cell activation and a pro-inflammatory milieu able to shape immune responses and limit the functionality of B-cell precursors.

ABCs are present at low levels in healthy individuals, but they tend to increase with age [7]. ABCs have been reported to be expanded in viral infections and autoimmunity, including RA, systemic lupus erythematous (SLE), multiple sclerosis, and Sjögren’s syndrome (reviewed in [7]). Evidence from RA populations derived from long-standing RA patients [6,10], mostly attributed to chronic stimulation, and the relevance of ABCs beyond disease activity is unknown. A recent study using single-cell profiling and flow cytometry has revealed that ABCs can be found in atherosclerosis plaques and blood in human and animal models [11]. Functional analyses revealed that ABCs may link immunity with the development of atherosclerosis lesions [11]. However, the clinical relevance of these finding is unclear.

Taken together, it is tempting to speculate that altered ABCs may be a common hallmark between RA pathogenesis and CVD. Altered immune (humoral) responses have been demonstrated to predate clinical diagnosis in RA [12], so ABCs may exhibit early alterations along RA course, probably connected with CVD endpoints which may provide clinical added value and allow for earlier interventions. Therefore, the main aims of the present study were (i) to evaluate ABCs levels in the earliest stages of inflammatory arthritis and their potential associations with clinical and immunological parameters, and (ii) to evaluate the role of ABCs as biomarkers for risk stratification.

## MATERIAL AND METHODS

### Study participants

Early RA patients (2010 ACR/EULAR classification criteria) [13] were recruited from the early arthritic clinic of the Department of Rheumatology at Hospital Universitario Central de Asturias (HUCA). RA patients were recruited at disease onset, and therefore were not exposed to disease-modifying antirheumatic drugs (DMARDs) at the time of sampling. A complete clinical examination, including Disease Activity Score 28-joints (DAS28), Simplified Disease Activity Index (SDAI) and Health Assessment Questionnaire (HAQ) calculations, was carried out during the clinical appointment. Patients were prospectively followed at 6 and 12 months. Clinical management was performed in compliance with EULAR recommendations [14], and clinical response was assessed using the EULAR response criteria [15].

Individuals with clinically suspect arthralgia (CSA) [16] were recruited from the same clinic. A group of healthy controls (HC) were recruited among age- and sex-matched healthy individuals from the same population.

A fasting blood sample was collected from all individuals by venepuncture in EDTA-containing tubes. Blood samples were immediately transferred to the laboratory and processed within less than 2 hours. Serum samples were stored at -80°C until experimental procedures. Conventional blood biochemical (including CRP and ESR measurements) and lipid analyses were performed in all individuals. Traditional CV risk factors (dyslipemia, hypertension, diabetes, smoking habit and obesity) according to national guidelines were registered from the medical records. CV risk was assessed using the mSCORE [17] and patients were grouped in risk categories (0-1: low, 1-5: medium; and >5: high and very high) according to the ESC consensus [18].

The study was approved by the local institutional review board (Comité de Ética de la Investigación con Medicamentos del Principado de Asturias) in compliance with the Declaration of Helsinki (reference CEImPA 2021.126). All study subjects gave written informed consent.

### ABCs assessment

PBMCs were isolated by centrifugation (1900 rpm, 20Lminutes) on density gradients (Lymphosep, Biowest, Germany). PBMCs were stained with anti-CD19 PerCP-Cy5,5 (Immunostep, Spain), anti-CD21 phycoerythrin (PE, Immunostep) and anti-CD11c allophycocyanin (APC, Immunostep) for 30 minutes at 4° C. Then, cells were washed twice with PBS and acquired in a BD FACS Canto II flow cytometer, using FACS Diva 2.6 for analysis. After gating lymphocyte population, CD19-positive events were selected and analyzed in a CD11c vs. CD21 dot-plot. CD11c-positive and CD21-negative events within the CD19+ subset were considered as ABCs [19].

### Vascular imaging and functional assessments

Doppler ultrasound assessment was performed in the sonography laboratory (Department of Neurology, HUCA) online in B-mode using a Toshiba Aplio XG device (Toshiba American Medical Systems) by an experienced user as previously described [20]. The carotid intima-media thickness (cIMT) was measured, according to the “Mannheim Carotid Intima-Media Thickness Consensus (2004-2006)” [21]. Atherosclerotic plaque burden and plaque vulnerability were defined according to established definitions as previously described [20].

Vascular stiffness was measured by evaluating the diameter changes of the common carotid during an entire cardiac cycle. To this end, stiffness parameters (vascular strain (VS) and stiffness (VSf), vascular distensibility (VD) and pressure-strain elastic modulus (PSEM)) were computed [22].

### Measurement of serum cytokines

The serum levels of IL-6, TNF, IFNg, IL-1b, IL-23, IL-12 and IL-8 were measured by a predefined multiplex assay (Human Inflammation Panel 1, LEGENDplex, BioLegend) in a BD FACS Canto II flow cytometer, according to the protocol provided by the manufacturer. The detection limits were 1.5 pg/ml, 0.9 pg/ml, 1.3 pg/ml, 1.5 pg/ml, 1.8 pg/ml, 2.0 pg/ml and 2.0 pg/ml, respectively.

APRIL, BAFF and IL-21 serum levels were quantified using ELISA kits (Invitrogen, ThermoFisher) in accordance with the manufacturer’s instructions. Detection limits for these cytokines were 0.40 ng/ml, 0.13 ng/ml and 20 pg/ml, respectively.

### Proteomic analysis

Serum proteomics were carried out by means of a high-throughput analysis using the Proximity Extension Assay provided by Olink (Olink Bioscience, Sweden) [23]. A pre-defined panel of 92 protein hits related to CVD (CV Panel II) was performed.

### Statistical analyses

Variables were summarized as median (interquartile range), mean±standard deviation, or as n(%), depending on their distribution. Differences among groups were evaluated by Mann-Withney U, Kruskal-Wallis or χ2 tests, as appropriate. Corrections for multiple comparison tests were performed by Dunn-Bonferroni. Correlations were assessed by Spearman ranks’ tests. The associations between ABCs counts and atherosclerosis occurrence were analyzed by logistic regression. Adjusted OR and 95% confidence intervals (CI) were calculated. The discrimination ability was assessed using the area under the receiver operating characteristic curve (AUC ROC). ABCs levels were added to the mSCORE algorithm, by pre-defined categories according to the distribution of the ABCs levels observed in the HC. Using tertiles as cut-offs, a score was given as follows: 0 to T1: +0, T1 to T2: +1, and T2 to T3: +2 points. These points were added to the mSCORE values to generate the new algorithm (mSCORE + ABCs).

The performance of the classification of mSCORE and mSCORE + ABCs was analysed by classification measures, correlation, and goodness of fit. The Youden index to determine the optimal cut-offs with maximum accuracy. Differences between AUC ROC curves were compared by the De Long statistic.

Proteomic data were evaluated pathway enrichment analyses using ShinyGo [24] and KEGG [25]platforms to identify biological pathways. Protein-protein interactions were assessed by the STRING database [26]. To investigate common transcriptional regulatory networks, the TRRUST [27] database was used to predict transcription factors underlying protein networks.

A p-value <0.050 was considered as statistically significant. Statistical analyses were carried out under SPSS v. 27, R v.4.1.3 and GraphPad Prism 8.0.

## RESULTS

### ABCs levels during the earliest stages of inflammatory arthritis

The levels of circulating ABCs were measured in 58 early RA patients, 11 individuals with CSA and 33 age and sex-matched HC (Table 1) by flow cytometry (Figure 1A). Statistical analyses revealed an ABCs expansion in RA patients compared to HC within the CD19+ total population (Figure 1B), and equivalent findings were observed within the PBMC compartment. Of note, a notable heterogeneity was observed within the RA group. Moreover, individuals with CSA showed numerically higher levels of ABC compared to the HC group, although statistical significance was not reached (Figure 1B). Similar findings were retrieved when absolute numbers were computed (Figure 1C).

**Table 1:**
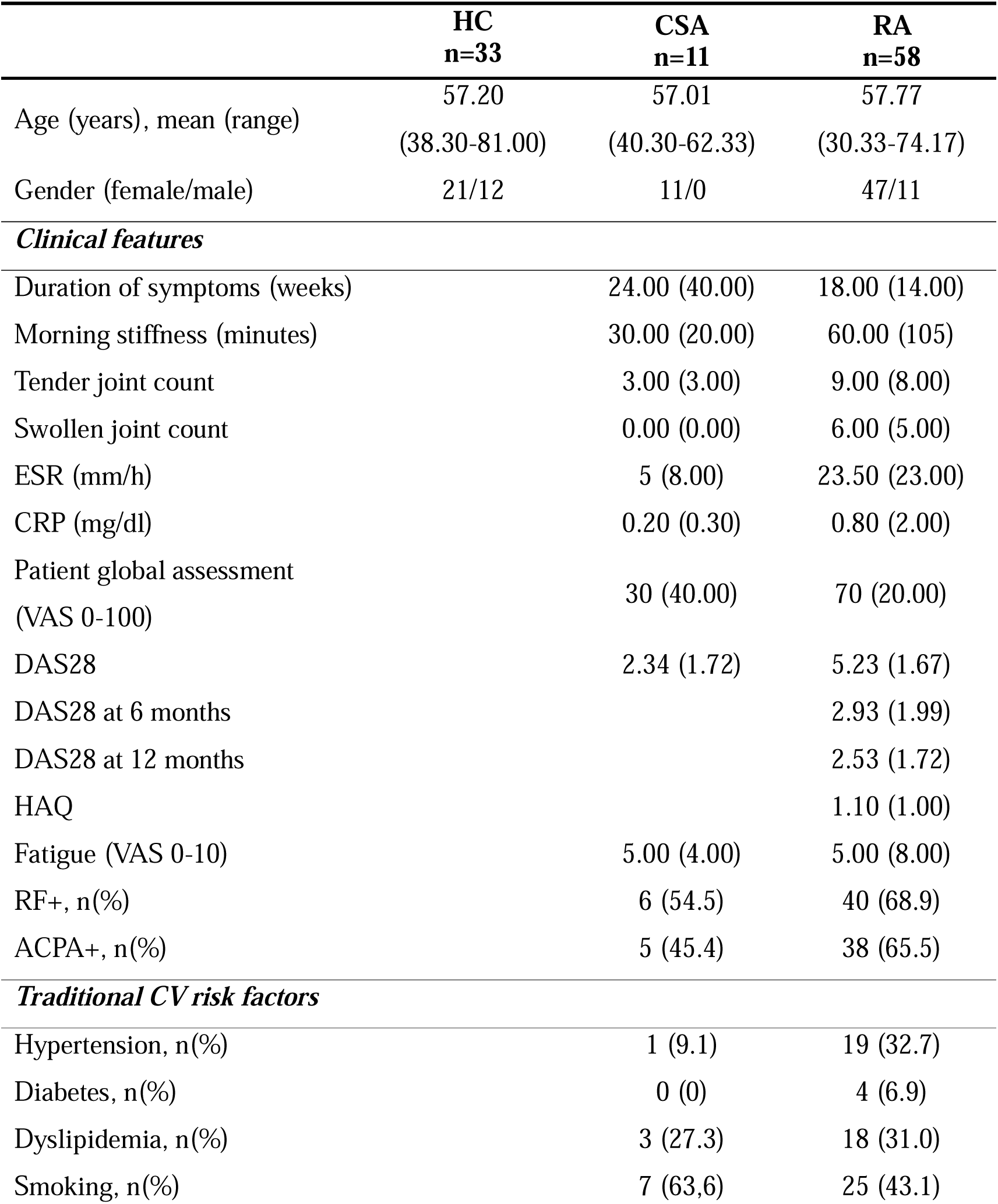
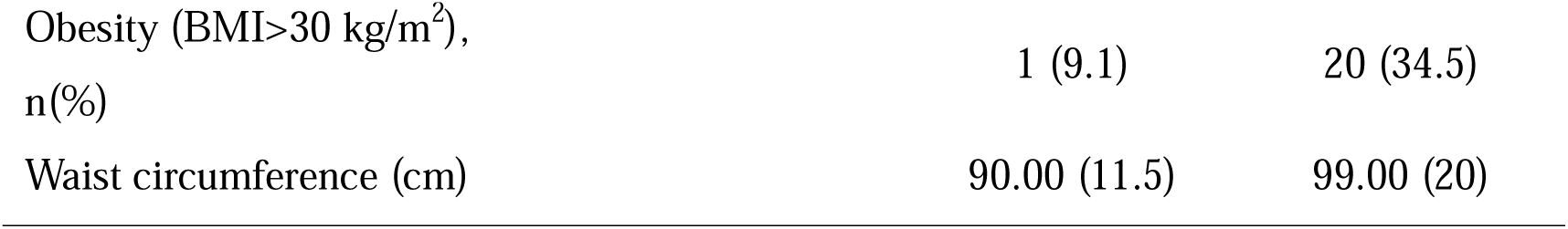
Demographic and clinical features of study participants. Demographic and clinical features of study participants were summarized. Variables were expressed as median (interquartile range), mean±SD or n(%), unless otherwise stated, according to the distribution of the variables.

**Figure 1:**
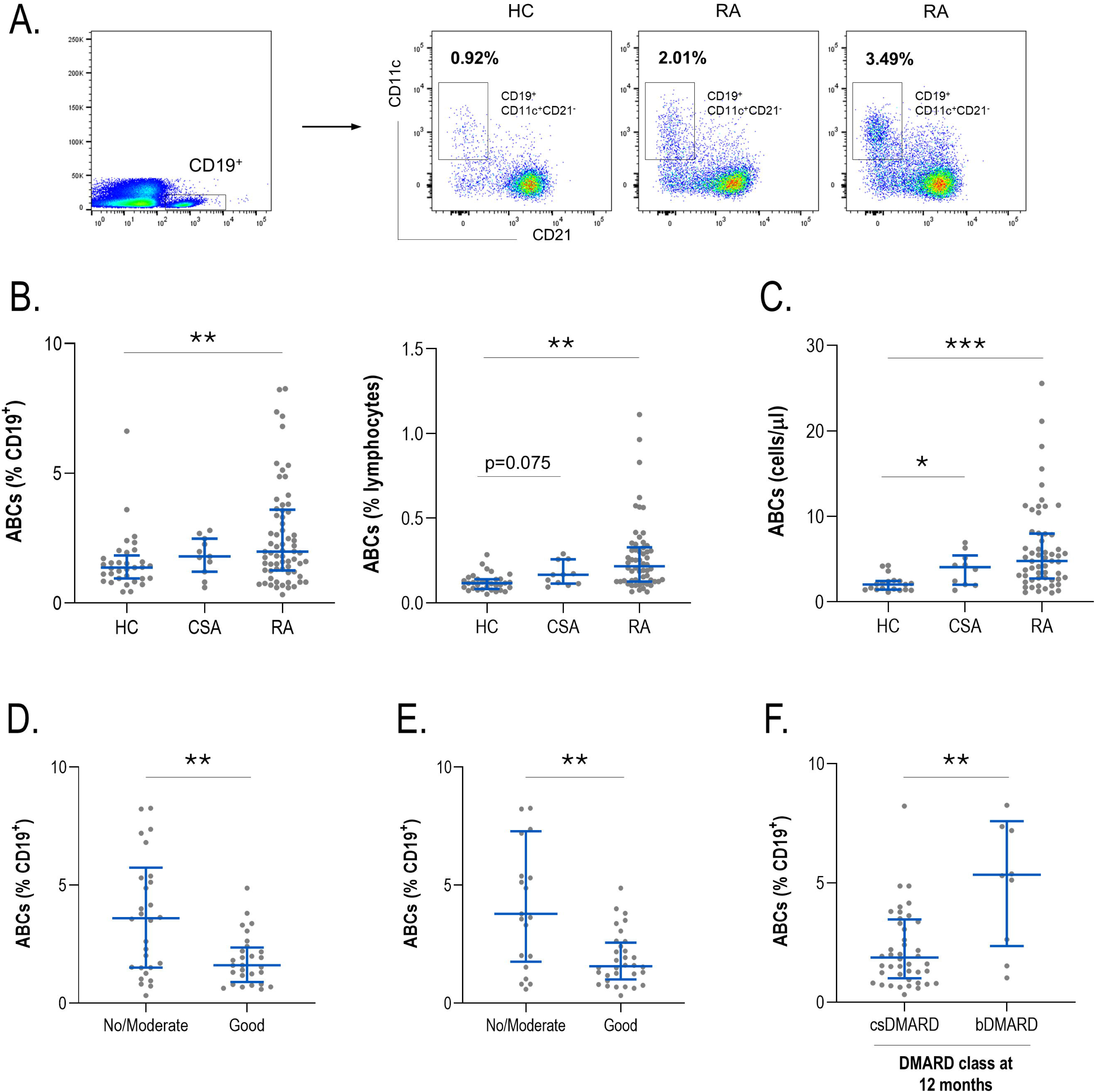
Analysis of ABCs levels across study groups. (A) Circulating ABCs levels were identified by flow cytometry. Representative dot plots from one HC and two RA patients are shown. ABCs frequency within CD19+ cells (A) or lymphocytes (B) were compared between HC, CSA individuals and RA patients. (C) Absolute levels of ABCs were computed and analyzed. The levels of ABCs were compared according to clinical response at 6 and 12 months (D-E), or according to treatment strategies at 12 months (F). Each dot represents one individual. Bars represent 25th percentile (lower), median and 75th percentile (upper). Differences were assessed by Kruskal-Wallis tests with Dunn-Bonferroni post-hoc tests. The p-values from the latter were indicated as follows: * p<0.050, ** p<0.010 and *** p<0.001.

The frequency of ABCs was unrelated to clinical features such as DAS28 (r=0.053, p=0.695), CDAI (r=0.002, p=0.988), HAQ (r=-0.318, p=0.341), symptom duration (r=0.159, p=0.259), or morning stiffness (r=-0.021, p=0.874) in RA patients. Similar findings were observed with age (r=0.175, p=0.190). ABCs levels were not associated with the presence of RF (p=0.638) or ACPA (p=0.722), nor with total serum levels of antibodies (IgG: r=-0.083, p=0.586; IgM: r=0.279, p=0.064; IgA: r=-0.027, p=0.858). Interestingly, lower levels of ABCs at baseline were observed in RA patients experiencing a good response to csDMARDs at 6 months (n=28) compared to those exhibiting no or moderate responses (n=30) (Figure 1D). Equivalent results were observed at 12 months (Figure 1E). In fact, patients switched to bDMARDs at 12 months due to clinical efficacy (n=9) showed higher ABCs frequency at baseline compared to those staying in the csDMARD treatment (Figure 1F).

Furthermore, correlation analyses revealed that ABCs were positively associated with a number of pro-inflammatory cytokines including IFNg, TNF, IL-6 and IL-21 in RA patients (Supplementary Table 1). Of note, no associations were observed with the levels of B-cell related factors such as BAFF or APRIL.

Taken together, these results suggest an ABCs expansion during the earliest stages of arthritis, which may be unrelated to clinical features but connected to treatment outcomes and certain inflammatory pathways.

### ABCs levels and subclinical CVD in early RA

Next, the associations between circulating ABCs levels and subclinical CVD outcomes were assessed in RA.

Our analyses revealed that patients with atherosclerosis plaques (n=33) exhibited higher ABCs levels within the CD19+ population compared to their atherosclerosis-free counterparts (n=21) (3.50 (3.53) vs 1.52 (1.42)%, p=0.006). Equivalent results were obtained within the total PBMC compartment (p=0.004). Interestingly, stronger associations between ABCs levels and pro-inflammatory cytokines were observed in patients with atherosclerosis (IL6: r=0.494, p=0.003; TNF: r=0.402, p=0.021; IFNg: r=0.509, p=0.002; IL-21: r=0.497, p=0.003), while being absent in those without atherosclerosis (all p>0.050). Furthermore, ABCs expansion was also associated with plaque number (Table 2). No associations with cIMT or vascular stiffness parameters were retrieved (Table 2). Similarly, ABCs frequency was unrelated to hypertension (p=0.193), dyslipidemia (p=0.619), smoking (p=0.772), diabetes (p=0.117), or obesity (p=0.361). Regression analyses revealed that ABCs counts were independent predictors of atherosclerosis occurrence in univariate or multivariate models, after adjusting for traditional CV risk factors (Table 3). Equivalent results were obtained when atherosclerosis plaque number was modeled in fully adjusted multivariate negative binomial regression analyses (ABCs: 1.734 [1.093 – 2.752], p=0.019), hence confirming the independent association between ABCs levels and atherosclerosis presence and extent.

**Table 2:** Associations between ABCs levels and subclinical CVD. Associations between ABCs levels and subclinical CVD outcomes were analyzed by Spearman ranks’ tests or Mann-Withney U tests in RA. Coefficients (r) and p-values, or p-values for the difference between groups are shown. Those reaching statistical significance are highlighted in bold.

|  |  |
| --- | --- |
| ***Subclinical atherosclerosis*** |  |
| Plaque presence | **p=0.006** |
| Plaque number | **r=0.346, p=0.010** |
| Plaque risk | p=0.265 |
| cIMT | r=0.139, p=0.322 |
| ***Vascular stiffness*** |  |
| VS | r=-0.165, p=0.336 |
| VD | r=-0.149, p=0.408 |
| VSf | r=0.222, p=0.214 |
| PSEM | r=0.149, p=0.408 |

**Table 3:** ABCs as predictor of atherosclerosis in RA. The role of ABCs levels as predictor of atherosclerosis occurrence in early RA patients was analysed by univariate and multivariate logistic regression models.

|  | **OR** | **95% CI** | **p-value** |
| --- | --- | --- | --- |
| ***Univariate*** |  |  |  |
| ABCs, per 1% | 1.260 | 1.051 – 1.510 | **0.012** |
| ***Multivariate*** |  |  |  |
| ABCs, per 1% | 1.284 | 1.035 – 1.593 | **0.023** |
| Sex, women | 0.055 | 0.003 – 1.185 | 0.055 |
| Age, per year | 5.331 | 1.558 – 18.241 | **0.008** |
| Smoking, yes | 6.448 | 1.049 – 39.646 | **0.044** |
| Hypertension, yes | 0.747 | 0.122 – 4.592 | 0.753 |
| Diabetes, yes | 0.001 | 0.000 – 0.001 | 0.999 |
| Dyslipidemia, yes | 0.804 | 0.145 – 4.462 | 0.803 |

ABCs alone were able to discriminate between patients with and withouth atherosclerosis (AUC ROC [95% CI], p: 0.733 [0.593 – 0.873], p=0.006). When patients were stratified according to mSCORE risk groups, this effect was mantained in the low-risk group (mSCORE<5, n=43) (0.716 [0.553 – 0.883], p=0.015). Therefore, we evaluated if ABCs levels could be useful in risk stratification. To this end, ABCs tertiles were added to the mSCORE (mSCORE + ABCs). Comparative analyses revealed that adding ABCs strata to the mSCORE improved the identification of RA patients with atherosclerosis into more realistic risk categories (Figure 2A). This further demonstrated by a better discrimination capacity (difference between areas = 0.149 [0.043–0.256], p=0.006), and improved classification metrics (sensitivity, percentage of patients correctly classified, and Matthews Correlation coefficient) (Figure 2B). NRI confirmed a better patient reclassification, with a higher effect for those presenting atherosclerosis and a marginal to no effect in the non-event group (Figure 2B). Finally, both a higher Youden index and a more realistic optimal cut-off for stratification (closer to 5) was achieved in the mSCORE+ABC classification compared with the mSCORE alone (Figure 2B).

**Figure 2:**
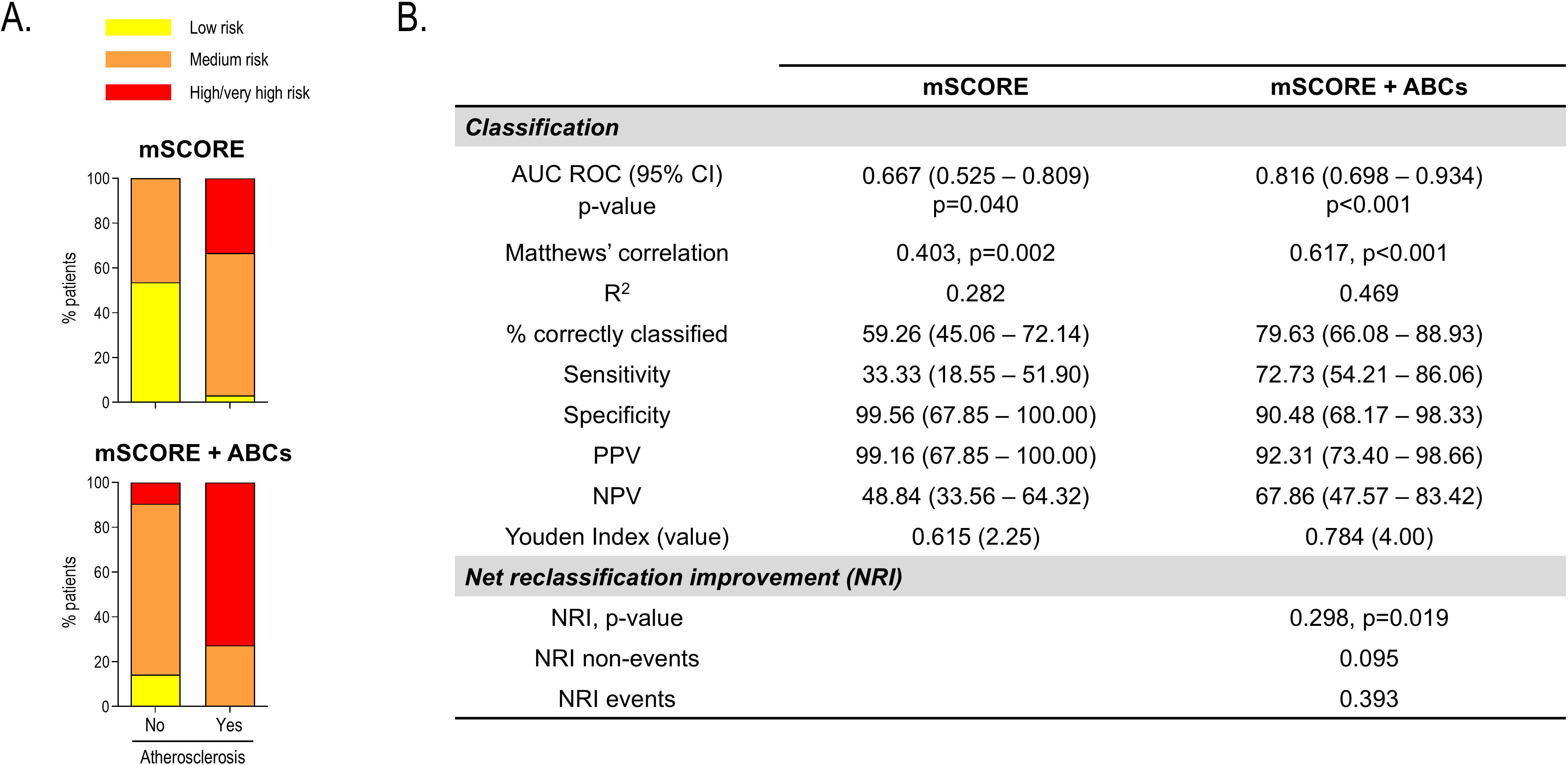
ABCs improved risk stratification in early RA. (A) Adding ABCs strata to existing algorithm (mSCORE) allowed risk stratification into more realistic risk categories, especially in patients with atherosclerosis. (B) Classification, calibration metrics and NRI statistics demonstrated an added value of ABS in improving risk stratification.

These results suggest that ABC expansion may be independently linked with atherosclerosis, also exhibiting a potential incremental value over existing algorithms for risk stratification.

### ABCs and proteomic signatures

Next, in order to get a more functional insight into the mechanisms behind ABCs expansion, the associations between ABCs levels and serum proteomic profiles were evaluated in RA patients.

Univariate correlation analyses revealed that ABCs counts were associated with a total of 11 protein hits (Figure 3A) (Supplementary Table 2), mostly exhibiting positive correlations. These proteins exhibited a high protein-protein interaction enrichment (p<3.3·10^-10^) (Figure 3B) using the STRING platform. Pathway annotation using KEGG mapper identified a number of relevant pathways (Figure 3C), mostly related to B-cell activation, T-B cell collaboration, T-cell dependent B-cell activation, T-cell polarization, and macrophages/foam cell differentiation. Pathway analysis using ShinyGO yielded other functional pathways including “regulation of lipid storage”, “leukocyte chemotaxis”, “mononuclear cell migration”, “cellular chemotaxis” or “cytokine-mediated signalling pathway”. Finally, analyses from the TRRUST database identified 10 candidate transcription factors shared for these proteins (Supplementary Table 3), hence reinforcing the occurrence of common expression programmes linked to these proteomic signatures.

**Figure 3:**
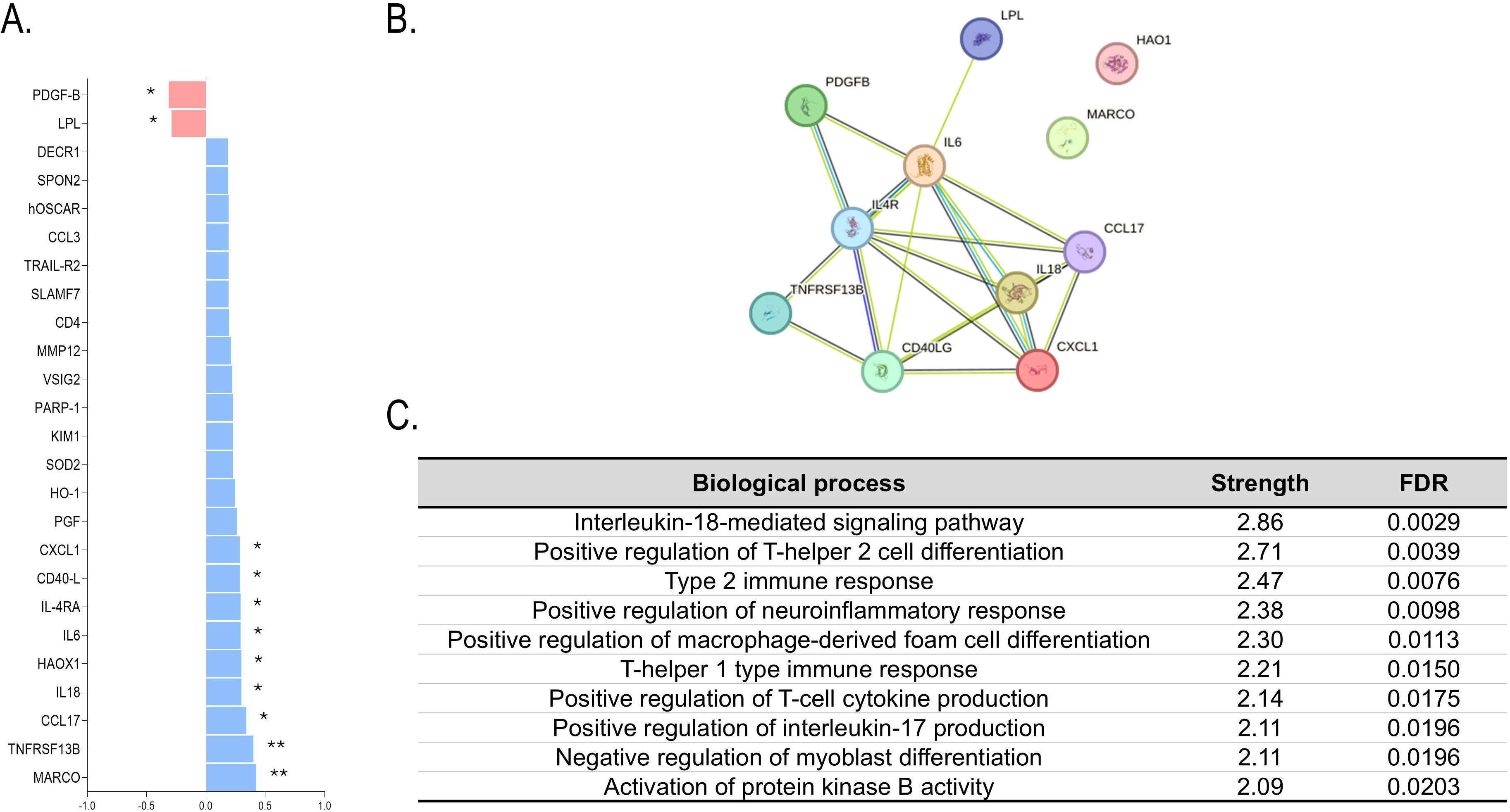
ABCs were related to proteomic signatures. (A) ABCs levels were associated with a number of protein hits in univariate correlation analyses. * p<0.050, ** p<0.010 (B) Protein-protein interactions among protein species found to be associated ABCs in early RA depicted in a network graph (STRING platform). (C) Functional classification of the proteomic species into biological pathways (top 10) retrieved by the STRING platform. Strength and enrichment FDR is indicated for each annotated pathway.

Taken together, all these findings suggest that ABCs expansion in RA patients is connected to specific protein signatures involved in B-cell activation, T-cell responses, and cellular pathways related to atherosclerosis development.

## DISCUSSION

Humoral responses have received less attention as the missing link between rheumatic diseases and CVD. However, the role of ABCs has emerged in recent years in several contexts such as autoimmunity and infection. The results herein presented demonstrated an early expansion of ABCs in the systemic compartment, which was connected with inflammatory circuits and atherosclerosis burden. To the best of our knowledge, this is the first characterization of ABCs during the earliest stages of inflammatory arthritis and their potential links with comorbidities in this condition.

A major breakthrough from our study was the characterization of the elevated ABCs levels during the earliest stages of inflammatory arthritis, including a subtle increase in individuals with CSA. Although expanded ABCs subsets have been reported in RA populations by other groups [19,28], whether this occurred at disease onset or if they accumulate along disease course due to senescence under chronic inflammation remained unknown. This is of special interest from pathogenic and therapeutic standpoints, as the links between altered ABCs levels and clinical outcomes in RA are yet to be established. Our findings demonstrate that ABCs expansion is an early event along arthritis course, independent of disease duration and drug exposure. These results align with evidence from animal studies, which show that autoimmune-prone mice strains exhibit higher circulating ABCs compared to their wild-type counterparts [19]. Furthermore, in the collagen-induce arthritis model, ABCs were detected in inguinal lymph nodes [29]. Interestingly, B-cell disturbances have been observed in human inguinal lymph nodes as well along early arthritis [30], also in relation to the T-cell compartment [31]. Taken together, these lines of evidence seem to suggest that ABCs expansion in RA predates clinical onset and thus its origin may be related to early immune dysregulation phenomena. A recent review has coined a ‘common triumvirate’ of stimuli for ABCs pathogenic generation, which includes BCR signals, TLR ligation and type-I inflammatory cytokine milieu [9]. It comes with no surprise that all these three events have been related to RA occurrence and development [3,32]. Importantly, TLR-7/9 over-activated signalling has been observed before the clinical onset in first-degree relatives of RA patients [33]. Moreover, these results also add to the field of senescence in rheumatic conditions [34]. Contemporary literature has pointed to an early and pronounced senescence phenomenon in patients with rheumatic conditions [35,36], which cannot be solely accounted by age, thus suggesting a gap between calendar and biological age. Whereas higher evidence is available for the T-cell compartment, B-cell subsets have been neglected. The findings herein presented align with this notion and may open new avenues to characterize (immune)-senescence in RA.

Our analyses revealed that ABCs frequencies were unrelated to clinical features, including autoantibody levels. This finding may not only indicate that ABCs expansion could be considered as a common hallmark on RA pathogenesis, but also that ABCs could be mainly involved in mechanisms other than antibody production. Actually, our results unveiled interesting associations between ABCs counts and the levels of certain proinflammatory cytokines. No associations were observed with B-cell-related factors, which is in line with the BAFF independency described elsewhere [37]. However, strong associations were observed with IL-21 and IFNg, major drivers of T-bet expression on B-cells and ABCs differentiation (reviewed in [7]). In fact, bystander cytokine production, particularly of IFNg, may suffice to drive ABCs generation [8]. Furthermore, IL-6 and TNF were also positively correlated with ABCs levels, and evidence is supportive of their role on ABCs biology. [8,9] These findings are central in this scenario, as these mediators are key for T-cell polarization as well as central for RA immune-pathogenesis [3,32]. By interpreting these findings based on the existing literature, it is tempting to speculate that ABCs may play a crucial role in shifting T-cell activation in RA. Functional analyses from our proteomic platform do reinforce this hypothesis. Moreover, ABCs have been described as highly potent antigen-presenting cells [7–9], hence strengthening their contribution to T-cell activation. These findings warrant a more in-depth elucidation of the involvement of ABCs on cytokine networks in RA, which may support their role as therapeutic targets.

Once ABCs expansion is established in RA, it is imperative to evaluate their relevance from a clinical perspective. Remarkably, our study demonstrated a solid association between ABCs counts and subclinical CVD. ABCs were known to migrate to synovial tissues in RA, and to participate in the index manifestation of arthritis, that is, synovial inflammation and bone erosion in a cytokine-mediated loop with fibroblast-like synoviocytes [38]. However, the role of ABCs in comorbidities and disease manifestations beyond the synovium had been largely unexplored. Our study demonstrated a strong association between ABCs and atherosclerosis burden and extent in early RA patients. This finding adds to the hypothesis of shared mechanisms between synovial inflammation and CVD in the context of RA [39], shedding light into a potential new mediator. Furthermore, ABCs accumulation was also related to poor therapeutic outcomes, which are in turn associated with higher CV risk in several studies [40,41]. Despite solid epidemiological evidence, mechanistic insight underlying this effect was rather absent. Then, ABCs may be a potential missing link in the connection between treatment failure and enhanced CVD burden. In this regard, whether patients with increased ABCs may be candidates for rituximab treatment should be evaluated, in order to both alleviate disease activity and CV burden. Rituximab has been proven to be effective in this context [42], although evidence is inconclusive [41]. Different trajectories among ABC and DN B-cells have been reported, which may explain these discrepancies [43].

The independent association between ABCs counts and atherosclerosis is relevant in a twofold manner. On the one hand, these findings strengthen the role of immune dysregulation to account for the enhanced CV risk in RA. Although the involvement of humoral responses had been suggested by our group and others, mostly by the production of antibodies, the role of B-cell subsets themselves has received less attention. Due to the multifaceted mechanisms described for ABCs, these could be regarded as relevant mediators in the connection between B-cell responses and CVD burden. Actually, although ABCs expansion had been observed in patients with coronary artery disease in a recent report [44], no associations were found with the levels of IgG or IgM antibodies against a MDA-mimotope, thus again pointing to their participation in mechanisms other than antibody production. These notions were also supported by our previous findings on the associations with pro-inflammatory cytokines and the evidence on the role of ABCs in modulating T-cell responses. On the other hand, this study demonstrates that ABCs provided added value that improved risk stratification over conventional algorithms. The fact that this study is based on subclinical CV endpoints may facilitate its use thereafter as a biomarker to identify patients with high-risk profiles to guide preventive strategies to combat clinical CVD, in line with existing guidelines. Actually, the identification of novel biomarkers to assist in clinical practice has been largely pursued in the research agenda of this field [17].

In conclusion, ABCs are expanded in the systemic compartment during the earliest stages of inflammatory arthritis, unrelated to clinical features but associated with poor therapeutic outcomes upon csDMARDs treatment and pro-inflammatory milieu. ABCs counts were independent predictors of atherosclerosis burden and improved risk stratification over conventional algorithms. Taken together, these results shed new light into the role of ABCs on immune circuits underlying early RA and paved the ground for its use as a biomarker for CV risk management in routine care. Larger and long-term studies are needed to validate these findings as well as to elucidate incremental value, including cost-effectiveness. Furthermore, prospective studies evaluating ABCs fluctuations under different DMARDs are warranted, in order to gain understanding towards their protective and detrimental effects on CV outcomes [41], and how this may impact CVD outcomes in the long-term. These findings may be of interest beyond rheumatology, and they may be applicable in a number of scenarios where inflammation and immune dysregulation has been demonstrated to drive CVD burden, including inflammaging.

## Data Availability

All data produced in the present work are contained in the manuscript

## ACKNOWLEDGEMENTS

The authors would like to thank Ms. Marta García Boto (Department of Rheumatology, HUCA) for her assistance in the collection of blood samples, and the “Liga Reumatológica Asturiana” for their support.

