## Supplementary Material for "Age-associated B-cells are expanded in early arthritis linked to atherosclerosis and immune circuits - a potential role as a biomarker for risk stratification"

**Supplementary Tables**

**SUPPLEMENTARY TABLES**

**Supplementary Table 1: Associations between ABCs levels HDL and serum cytokines.** Associations between ABCs frequencies and serum levels of cytokines were analysed by Spearman’s rank tests in CSA and RA groups. Coefficients (r) and p-values are shown. Those reaching statistical significance are highlighted in bold.

|  | **CSA** | **RA** |
| --- | --- | --- |
| IL-6 | r=0.141  p=0.679 | **r=0.324**  **p=0.013** |
| TNF | r=-0.118  p=0.729 | **r=0.252**  **p=0.047** |
| IFNa | r=0.400  p=0.223 | r=0.031  p=0.817 |
| MIP1a | r=0.357  p=0.125 | r=0.089  p=0.756 |
| IFNg | r=0.028  p=0.936 | **r=0.355**  **p=0.006** |
| IL-1b | r=0.341  p=0.305 | r=0.176  p=0.186 |
| IL-33 | r=-0.091  p=0.790 | r=0.125  p=0.349 |
| IL-23 | r=-0.091  p=0.790 | r=0.138  p=0.301 |
| IL-18 | r=0.209  p=0.537 | r=0.125  p=0.352 |
| IL-17 | r=0.136  p=0.689 | r=0.012  p=0.931 |
| IL-12 | r=0.219  p=0.517 | r=0.126  p=0.347 |
| IL-10 | r=-0.009  p=0.979 | r=0.075  p=0.574 |
| IL-8 | r=-0.050  p=0.884 | r=0.096  p=0.474 |
| IL-21 | r=0.173  p=0.612 | **r=0.324**  **p=0.013** |
| BAFF | r=0.356  p=0.282 | r=-0.049  p=0.714 |
| APRIL | r=-0.145  p=0.670 | r=0.132  p=0.322 |

**Supplementary Table 2: List of proteins associated with ABCs levels in RA.** Serum proteins found to be associated with ABCs are listed along with their KEGG ortholog codes and most representative functions.

| **Protein** | **KEGG ortholog** | **Functions** |
| --- | --- | --- |
| TNFRSF13B | K05150 | Stimulation of B- and T-cell function and the regulation of humoral immunity |
| IL-6 | K05405 | Immune activation |
| IL-18 | K10030 | Immune activation |
| CXCL1 | K05505 | Neutrophil activation and migration; endothelial activation |
| LPL | K01059 | Triglyceride and TRL metabolism  Lipid clearance |
| IL-4RA | K05071 | Immune regulation |
| PDGF-B | K17386 | Cell proliferation, cell migration, survival and chemotaxis  Recruitment of vascular smooth muscle cells  Blood vessel development and wound healing |
| CD40-L | K03161 | Costimulates T-cell proliferation and cytokine production  Activation of NF-kappa-B  B-cell proliferation |
| HAOX1 | K11517 | Fatty acid metabolism |
| CCL17 | K21083 | T-cell chemotaxis  T-cell activation |
| MARCO | K13884 | Macrophage activation and differentiation  Phagocytosis |

**Supplementary Table 3: List of candidate transcription factors for the proteins analyzed.** Transcription factors identified to be shared candidate key regulators for the proteins analyzed using the TRRUST database.

| **Abbreviation** | **Transcription factor** | **# overlapped genes** | **p-value** | **FDR** |
| --- | --- | --- | --- | --- |
| RELA | v-rel reticuloendotheliosis viral oncogene homolog A (avian) | 5 | 4.26·10^-7^ | 2.22·10^-6^ |
| NFKB1 | nuclear factor of kappa light polypeptide gene enhancer in B-cells 1 | 5 | 4.40·10^-7^ | 2.22·10^-6^ |
| REL | v-rel reticuloendotheliosis viral oncogene | 2 | 7.09·10^-5^ | 0.0002 |
| FOXO1 | Forkhead box O1 | 2 | 8.47·10^-5^ | 0.0002 |
| SP1 | Sp1 transcription factor | 4 | 0.0001 | 0.0002 |
| JUND | jun D proto-oncogene | 2 | 0.0001 | 0.0002 |
| GATA3 | GATA binding protein 3 | 2 | 0.0002 | 0.0002 |
| PPARA | peroxisome proliferator-activated receptor alpha | 2 | 0.0002 | 0.0002 |
| STAT1 | signal transducer and activator of transcription 1 | 2 | 0.0010 | 0.0011 |
| EGR1 | early growth response 1 | 2 | 0.0011 | 0.0011 |
